# The Outcome Impact of Early vs Late HFNC Oxygen Therapy in Elderly Patients with COVID-19 and ARDS

**DOI:** 10.1101/2020.05.23.20111450

**Authors:** Liehua Deng, Shaoqing Lei, Fang Jiang, David A. Lubarsky, Liangqing Zhang, Danyong Liu, Conghua Han, Dunrong Zhou, Zheng Wang, Xiaocong Sun, Yuanli Zhang, Chi Wai Cheung, Sheng Wang, Zhong-yuan Xia, Richard L Applegate, Hong Liu, Jing Tang, Zhengyuan Xia

**Author notes:** Drs. Deng, Lei and Jiang contributed equally as co-first authors. Drs. H. Liu, J. Tang and Z. Xia contributed equally as co-last authors. Joint corresponding authors Dr. Zhengyuan Xia, Department of anesthesiology, the University of Hong Kong, Hong Kong, China; and Department of Anesthesiology, Affiliated hospital of Guangdong Medical University, China, Dr. Jing Tang, Department of Anesthesiology, Affiliated hospital of Guangdong Medical University, China, Dr. Liehua Deng, Department of Critical Care Medicine of affiliated hospital of Guangdong Medical University, Guangdong, China.

## Abstract

Coronavirus disease-2019 (COVID-19) has rapidly spread worldwide. High-flow nasal cannula therapy (HFNC) is a major oxygen supporting therapy for severely ill patients, but information regarding the timing of HFNC application is scarce, especially in elderly patients. We retrospectively analyzed the clinical data of 110 elderly patients (≥65 years) who received HFNC from Renmin Hospital of Wuhan University, People’s Hospital of Xiantao City and Chinese Medicine Hospital of Shishou City in Hubei Province, China, and from Affiliated Hospital of Guangdong Medical University, People’s Hospital of Yangjiang City, People’s Hospital of Maoming City in Guangdong Province, China.

Of the 110 patients, the median age was 71 years (IQR, 68-78) and 59.1% was male. Thirty-eight patients received HFNC when 200 mmHg < PO_2_/FiO_2_ ≤ 300 mmHg (early HFNC group), and 72 patients received HFNC treatment when 100 mmHg < PaO_2_/FiO_2_ ≤ 200 mmHg (late HFNC group). Compared with the late HFNC group, patients in the early HFNC group had a lower likelihood of developing severe ARDS, longer time from illness onset to severe ARDS and shorter duration of viral shedding after illness onset, as well as shorter lengths of ICU and hospital stay. Twenty-four patients died during hospitalization, of whom 22 deaths (30.6%) were in the late HFNC group and 2(5.3%) in the early HFNC group. It is concluded that the Prognosis was better in severely ill elderly patients with COVID-19 receiving early compared to late HFNC. This suggests HFNC could be considered early in this disease process.

## Introduction

Coronavirus disease-2019 (COVID-19) caused by severe acute respiratory syndrome coronavirus 2 (SARS-CoV-2), was first reported in December 2019 in Wuhan, Hubei, China, but has rapidly spread worldwide.[1] Since initial detection of the virus, more than 2,626,000 cases of COVID-19 have been confirmed worldwide, with more than 181,000 deaths as of April 24, 2020. COVID-19 is more likely to affect elderly patients with comorbidities, and can result in severe or fatal respiratory diseases such as acute respiratory distress syndrome (ARDS).[2] Initial reports from China[3], Italy[4] and the United States[5] suggested high mortality for elderly and critically ill patients with COVID-19. No specific therapeutic agents or vaccines for COVID-19 are available, although several antiviral medications such as remdesivir and favipiravir are under investigation. [6’ 7]

Patients with severe illness may develop dyspnea and hypoxemia within one week after the onset of COVID-19 and may quickly progress to ARDS, [2] a major cause of death in patients with COVID-19.[8] Thus, respiratory support and intensive care management are vital to saving lives. Reports showed that both conventional oxygen therapy and non-invasive ventilation (NIV), such as NIPPV (non-invasive positive pressure ventilation), are commonly used in COVID-19 patients to improve oxygenation and reduce the possibility of intubation.[9’ 10] However, the risk of airborne transmission with NIPPV is a major concern, while that for high flow nasal cannula therapy (HFNC) is judged minimal.[11] A guideline for the management of critically ill adults with COVID-19 published in JAMA March 26, 2020 recommended the use of HFNC relative to NIPPV in the circumstance of acute hypoxemic respiratory failure despite conventional oxygen therapy.[11] However, evidence is lacking regarding optimal timing to apply HFNC. In this study, we report the clinical characteristics of elderly (≥65 years) patients with COVID-19 who developed ARDS on or shortly after admission and compared outcomes of patients who started HFNC at different stages of ARDS.

## Methods

### Study design and participants

This retrospective cohort study included elderly patients (≥ 65 years) from Renmin Hospital of Wuhan University, People’s Hospital of Xiantao City and Chinese Medicine Hospital of Shishou City in Hubei Province, China, and from Affiliated Hospital of Guangdong Medical University, People’s Hospital of Yangjiang City, People’s Hospital of Maoming City in Guangdong Province, China. All elderly patients who were diagnosed with severe COVID-19 according to WHO interim guidance, and those who were treated with HFNC between January 14, 2020 (when the first patients were admitted in these hospitals) and March 5, 2020, were included in the present study. Due to the lack of understanding of this new disease, the timing of HFNC treatment was uncertain. In this retrospective study, of 110 HFNC-treated patients, 38 received HFNC treatment when 200 mmHg < PO_2_/FiO_2_ ≤ 300 mmHg (early HFNC group), while 72 were first treated with conventional oxygen therapies (e.g., low flow nasal catheter ventilation) and then HFNC when 100 mmHg < PO_2_/FiO_2_ ≤ 200 mmHg (late HFNC group). HFNC was started from low levels and gradually titrated to 60 L/min for patients without obvious complaint of chest distress or shortness of breath. However, for patients who were short of breath (e.g., respiratory rate >30/min) the flow rates were commenced at 60 L/min. The goal of oxygen therapy was to maintain the oxygen saturation (SpO_2_) at 93%-96%, which is in keeping with the recent guideline recommendation of a reasonable SpO2 range of 92-96% for patients receiving oxygen.[12]

This study was reviewed and approved by the Medical Ethical Committee of participating institutes (PJ2020-005), and the informed consent was waived by the Medical Ethical Committee.

### Data Collection

Patients’ medical records were reviewed and epidemiological, clinical, laboratory, and radiological characteristics and treatment and outcomes data were obtained with data collection forms. Two research investigators (LD and SL) independently reviewed the data collection forms to verify accuracy.

We collected data on age, sex, exposure history, comorbidities (e.g., hypertension, diabetes, malignancy, cardiovascular disease, cerebrovascular disease, chronic pulmonary disease, chronic kidney disease), chest CT images, signs and symptoms (e.g., fever, fatigue, dry cough, dyspnea), time of first symptom to dyspnea, ARDS and ICU admission, vital signs (heart rate, respiratory rate, blood pressure) and laboratory values (e.g., white blood cell count, neutrophil count, lymphocyte count, procalcitonin concentration, arterial blood gas analysis, fraction of inspired oxygen (FiO_2_), partial pressure of arterial oxygen (PaO_2_), and lactate concentration) on hospital admission and disease progression, treatments (e.g., oxygen support, antiviral therapy, antibiotic therapy, glucocorticoids, immunoglobulin), complications (e.g., septic shock, ARDS, secondary infection), and discharge/death. The numbers of patients requiring mechanical ventilation, the numbers of patients requiring FiO_2_ = 100%, > 80% and > 60% for more than 72 hours continuously, length of ICU stay, and length of stay (LOS) were also collected.

### Outcomes

The primary outcome was in-hospital mortality. Secondary outcomes included incidence of severe ARDS, the numbers of ICU admission and patients requiring mechanical ventilation. The ICU admission standard is patients require invasive mechanical ventilation, or have shock or other organ failure that need ICU monitoring and treatment.[13’ 14] ARDS was defined as acute onset hypoxemia (PaO_2_/FiO_2_: mild ARDS, > 200 to ≤ 300 mmHg; moderate ARDS, >100 to ≤200 mmHg; severe ARDS, ≤100 mmHg) with bilateral pulmonary opacities on chest imaging not fully explained by other disease according to the Berlin definition.[15] Secondary infection was defined when patients showed clinical symptoms or signs of bacteremia and a positive culture of a new pathogen obtained from sputum or blood samples after admission. [13] Acute cardiac injury was identified when the hypersensitive troponin I and creatine kinase-MB were above the 99% upper reference limit or new abnormalities were shown in electrocardiography and echocardiography.[13] Acute kidney injury (AKI) was defined according to KDIGO criteria.[16]

### Statistical analysis

Continuous variables were presented as mean with standard deviation (SD) when normally distributed and compared by independent sample *t* test, or expressed as median with interquartile range (IQR) if non-normally distributed and compared by Mann-Whitney U test. Categorical variables were expressed as n (%) and compared by Pearson’s chi-square or Fisher’s exact test between early HFNC and late HFNC groups. A two-sided *α* of less than 0.05 was considered statistically significant. All statistical analyses were performed with the SPSS (version 25) software.

## Results

A total of 638 elderly patients (≥ 65 years) with confirmed SARS-CoV-2 infection were admitted to participating hospitals during the defined study time period. Of these, 502 patients who did not receive HFNC treatment were excluded, as were 19 patients due to missing key information in their medical records, and 7 patients who had cardiac arrest within 24 hours after admission. Thus, a total of 110 patients were included in our study. Of these 110 patients, 38 patients received early HFNC treatment, and 72 patients received late HFNC treatment.

The median age of the 110 patients was 71 (IQR 68-78; range 65 to 89) years, and most (65[59.1%)]) were male (Table 1). Eighty-seven (79.1%) patients had underlying comorbidities, 1.3 comorbidities per patient on average. The most common comorbidities were hypertension (57 [51.8%]), cardiovascular disease (27 [24.5%]), chronic pulmonary disease (22 [20%]) and diabetes (20 [18.2%]). The most common symptoms on admission were fever (105 [95.5%]), cough (65 [59.1%]), weakness (23 [20.9%]), and sputum production (22 [20%]) Table 1). The most common abnormal laboratory findings were lymphocytopenia, increased C-reactive protein, and decreased CD3, CD4 and CD8 counts on hospital admission (Table 1). The overall median SPO_2_ was 95% (IQR 93-98%) on admission, and the median ratio of PaO_2_/FiO_2_ was 238 mmHg (IQR 221-277). There were no significant differences in admission SPO_2_ or PaO_2_/FiO_2_ between early and late HFNC groups. Both SpO_2_ and PaO_2_/FiO_2_ ratio at initiation of HFNC were higher and the time from admission to HFNC treatment was shorter in the early HFNC group (Table 1).

**Table 1.**
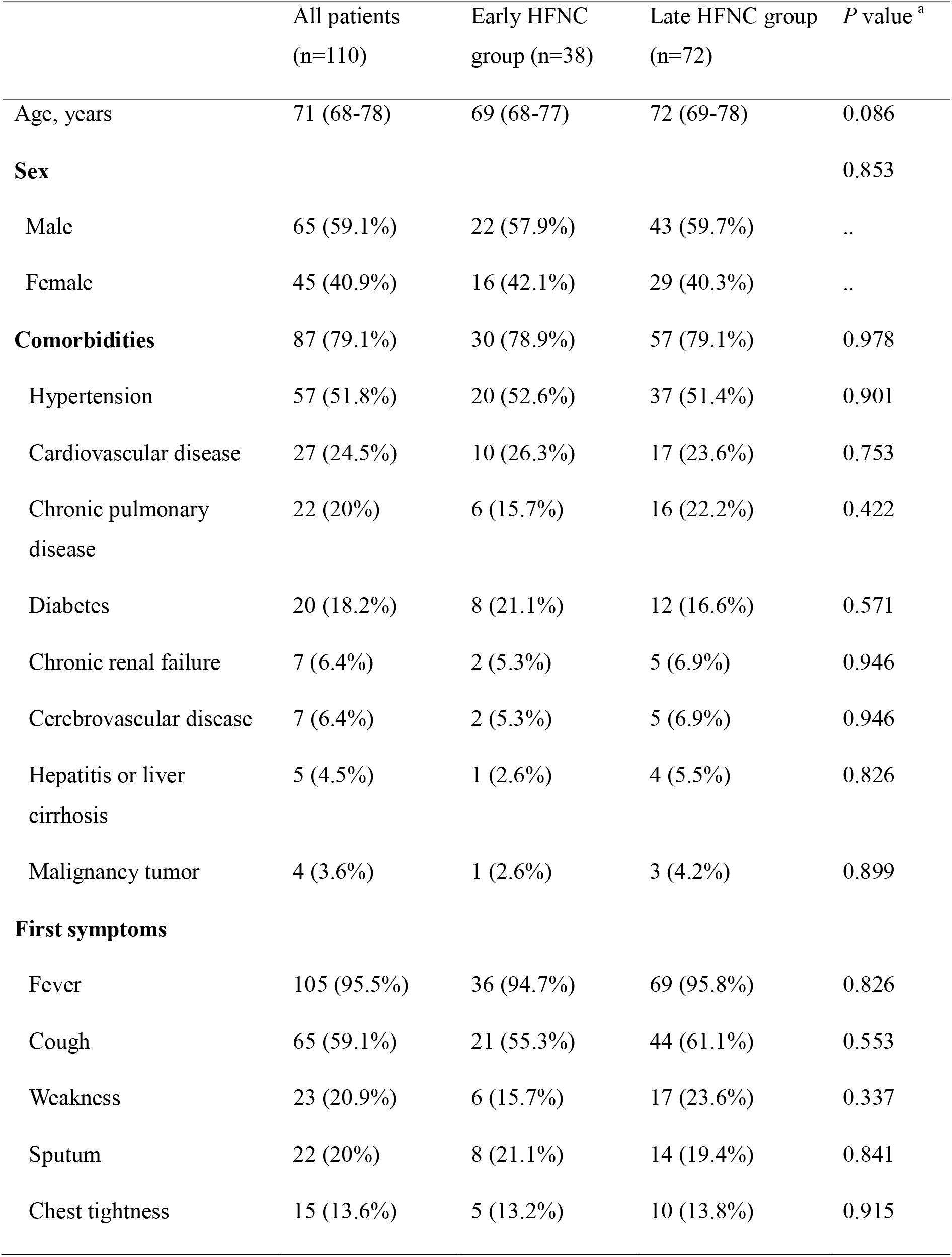
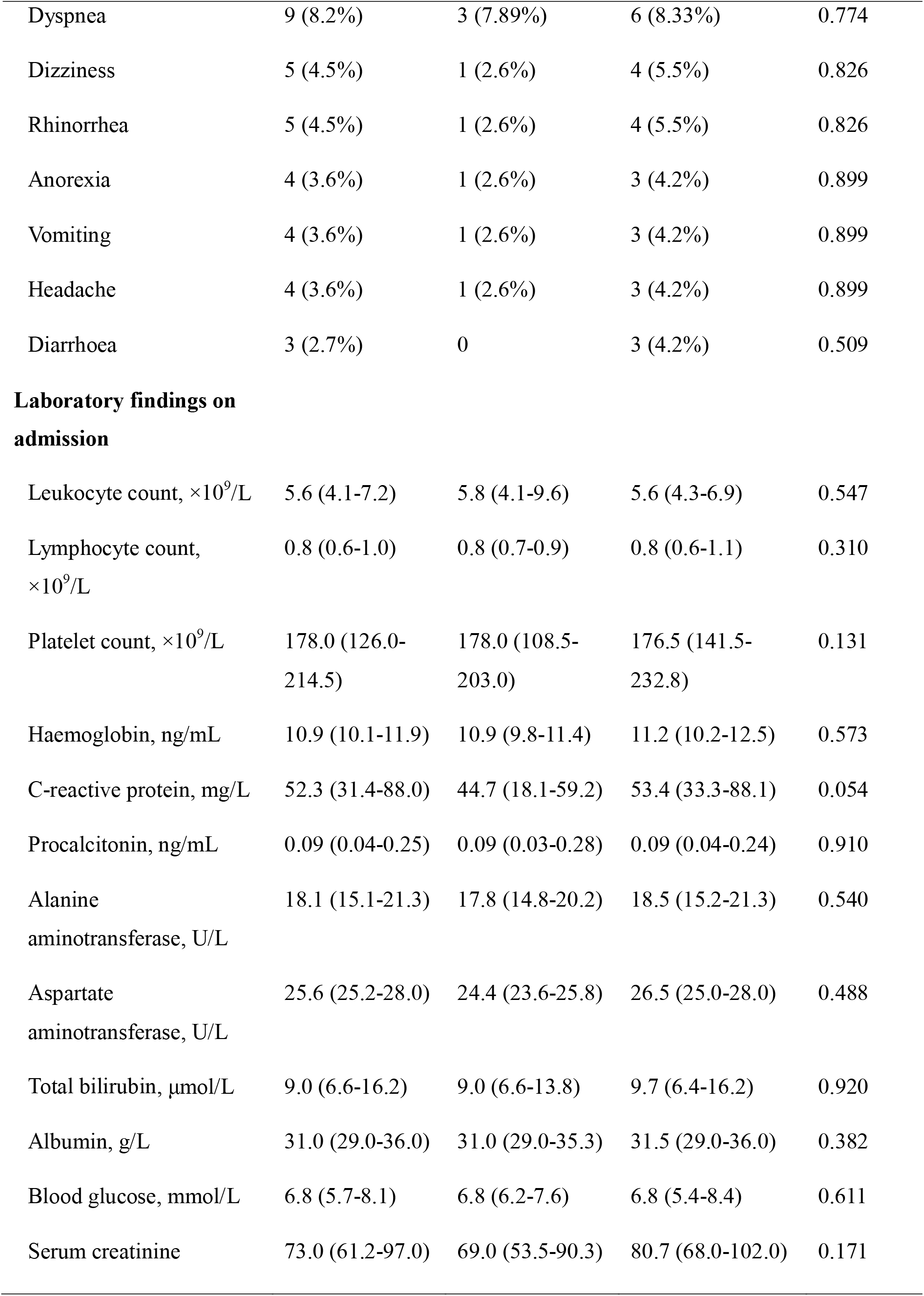
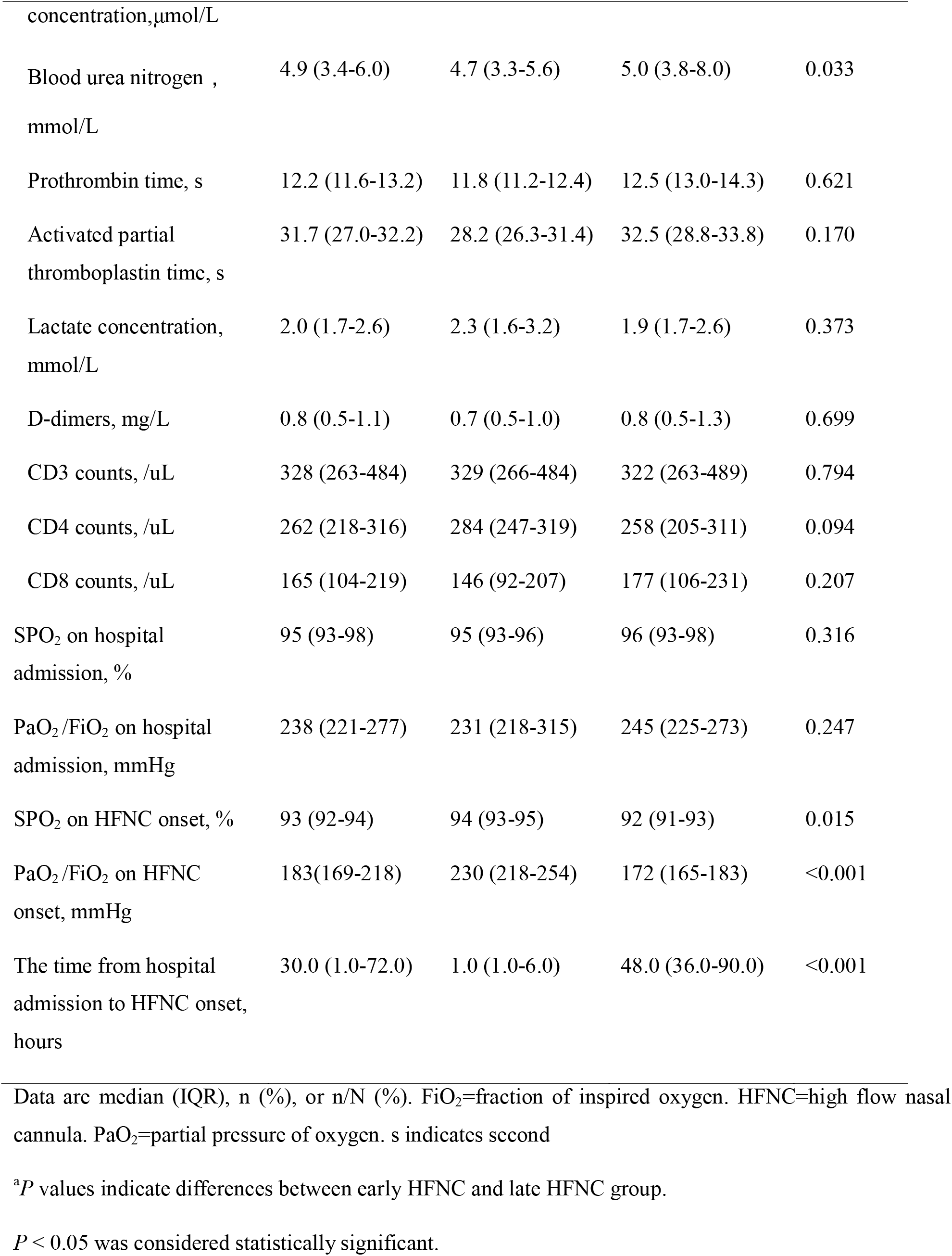
Demographic, clinical characteristics and laboratory findings of the study cohort

All patients showed bilateral lung involvement on chest CT scan at admission, including consolidation, ground-glass opacity, interstitial lesions, and exudative lesions (figure 1). Patients who received early HFNC had a lower likelihood of developing severe pneumonia, manifested as more than 50% increase in pneumonitis foci on chest CT scan during disease progression (Table 2).

**Figure 1.**
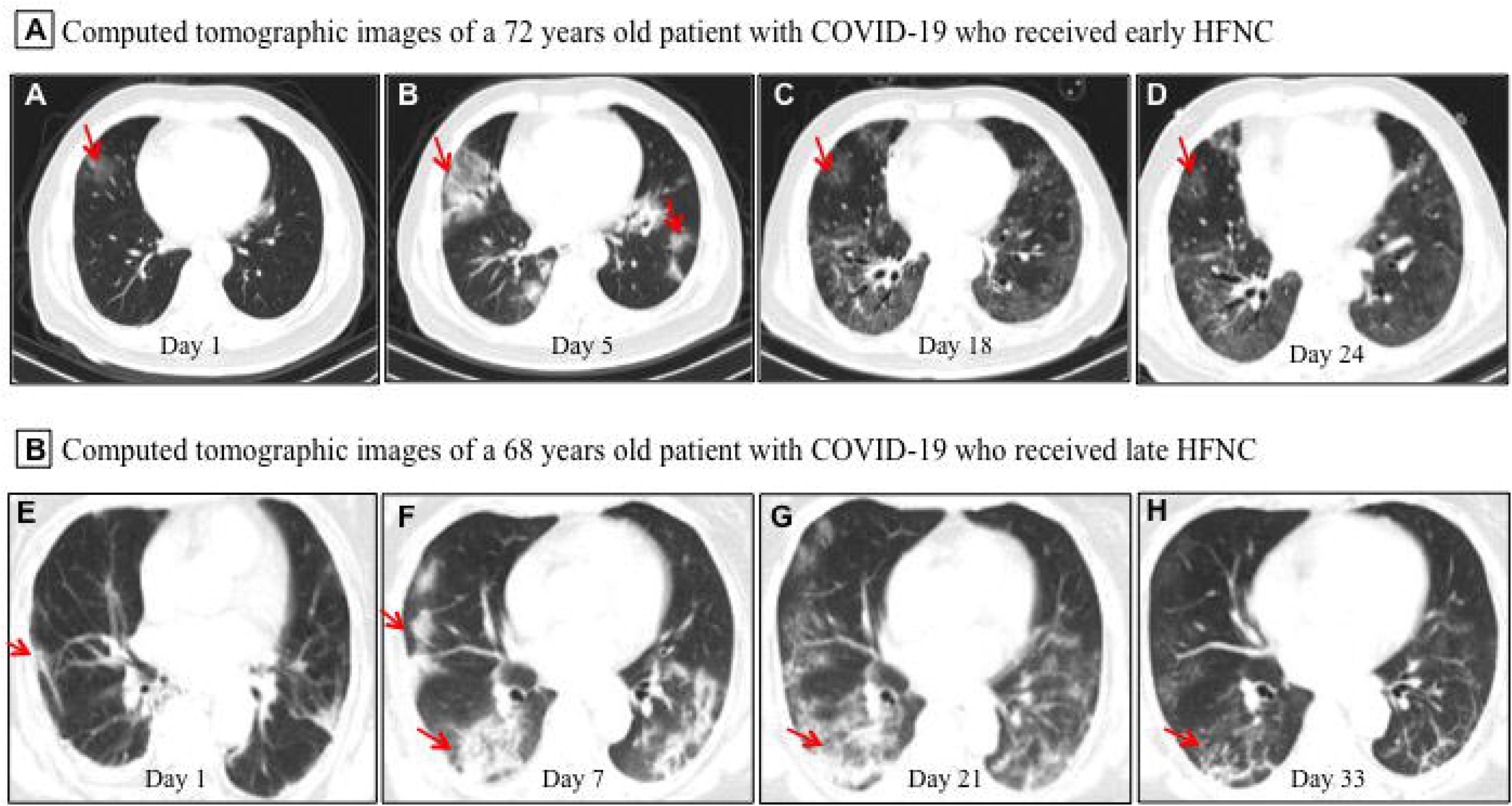
Representative chest computed tomographic images of elderly patients with COVID-19 who received early or late High-flow nasal cannula (HFNC). **A-D:** a 72 year old man with COVID-19 received early HFNC therapy when his PaO_2_/FiO_2_ was 256 mmHg. (A) Image obtained on day 1 showed small ground-glass opacity lesion (red arrow) in the middle lobe of the right lung; (B) image obtained on day 5 showed enlarged lesion in right lung and several small areas of new exudative lesions in outer basal segment of lower lobe of left lung; (C) image obtained on day 18 showed the lesion in the middle lobe of the right lung reduced more than 50%, and clear interstitial lesions were found in the lower lobes of both lungs; (D) image obtained day 24 showed the lesions were further reduced and became lighter in density (red arrow). HFNC was discontinued on day 23, and this patient was discharged on Day 26. **E-F:** a 68 year old man with COVID-19 received late HFNC oxygen therapy when his PaO_2_/FiO_2_ was 186 mmHg. (E) Image obtained on day 1 showed a few patchy exudative lesions and cord like fibrosis in bilateral lobes of both lungs (red arrow); (F) image obtained on day 7 showed original lesions were obviously increased, and parenchymal lesions (such as consolidation and air bronchogram) in the middle and lower lobes of right lung, as well as appearance of interstitial lesions in lower left lung; (G) image obtained on day 21 showed increased patchy exudative lesions and interstitial lesions with light density (a few reticular lung changes) in lower left lung; (F) image obtained on day 33 showed a few grid lung changes and subpleural lines in the right lower lobe. This patient required invasive mechanical ventilation on day 23, and died of cardiac arrest on day 36 after admission.

**Table 2.** Outcomes of the study cohort

|  | All patients<br>(n = 110) | Early HFNC<br>group (n = 38) | Late HFNC<br>group (n = 72) | P value <sup>a</sup> |
| --- | --- | --- | --- | --- |
| **Treatment** |  |  |  |  |
| Antiviral therapy | 100 (100%) | 38 (100%) | 72 (100%) | NA |
| Antibiotic therapy | 66 (60%) | 20 (52.6) | 46 (63.9) | 0.027 |
| Prone ventilation | 38 (34.5%) | 3 (7.9%) | 34 (47.2%) | <0.001 |
| Non-invasive mechanical ventilati | 24 (21.8%) | 6 (15.7%) | 18 (25%) | 0.266 |
| Invasive mechanical ventilation | 42 (38.2%) | 4 (10.5%) | 38 (52.7%) | <0.001 |
| ECMO | 7 (6.4%) | 1 (2.6%) | 6 (8.3%) | 0.450 |
| **Complications** |  |  |  |  |
| Secondary infection | 51 (46.3%) | 10 (26.3%) | 41 (56.9%) | 0.002 |
| Severe ARDS | 42 (38.2%) | 4 (10.5%) | 38 (38.9%) | 0.037 |
| Septic shock | 18 (16.4%) | 3 (7.8%) | 15 (20.8%) | 0.081 |
| Cardiovascular event | 13 (11.8%) | 2 (5.3%) | 11(15.3%) | 0.216 |
| Acute kidney injury | 6 (5.4%) | 1 (2.6%) | 5 (6.9%) | 0.613 |
| Cardiac arrest | 3 (2.7%) | 0 | 3 (4.2%) | 0.518 |
| Chest CT foci increasing ≥ 50% | 43 (39.1%) | 8 (21.1%) | 35 (48.6%) | 0.004 |
| The time from illness onset to<br>severe ARDS, days | 12 (11-15) | 15 (13-17) | 11 (9-13) | <0.001 |
| The time from illness onset to<br>ICU admission, days | 11 (8-14) | 14 (10-16) | 11 (8-13) | <0.001 |
| ICU admission | 62 (56.4%) | 10 (26.3%) | 52 (72.2%) | <0.001 |
| ICU stay $\geq 7$ days | 57 (51.8%) | 14 (36.8%) | 43 (59.7%) | 0.022 |
| Length of ICU stay, days | 17 (12-22) | 12 (10-15) | 18 (13-22) | <0.001 |
| Length of stay, days | 27 (16-32) | 16 (15-22) | 30 (27-33) | <0.001 |
| Duration of viral shedding after illness onset, days | 16 (13-21) | 12 (9-15) | 18 (15-25) | <0.001 |
| **FiO<sub>2</sub> during hospitalization** |  |  |  |  |
| 100% more than 72 hours continuously | 4 (3.6%) | 0 | 4 (5.5%) | 0.345 |
| $\geq 80\%$ more than 72 hours continuously | 23 (22.7%) | 3 (7.9%) | 20 (27.8%) | 0.015 |
| $\geq 60\%$ more than 72 hours continuously | 40 (36.4%) | 4 (10.5%) | 36 (50%) | <0.001 |
| **Prognosis** |  |  |  | 0.002 |
| Discharge | 86 (78.2%) | 36 (94.7%) | 50 (69.4%) | .. |
| Death | 24 (21.8%) | 2 (5.3%) | 22 (30.6%) | .. |

Data are median (IQR), n (%), or n/N (%). ECMO=extracorporeal membrane oxygenation. FiO<sub>2</sub>=fraction of inspired oxygen. HFNC=high flow nasal cannula. ICU=intensive care unit. PaO<sub>2</sub>=partial pressure of oxygen. NA=not applicable.
<sup>a</sup>*P* values indicate differences between early HFNC and late HFNC group.
*P* < 0.05 was considered statistically significant.

All patients received antiviral medications (lopinavir or ritonavir), and 66 (60%) patients also received antibiotics. Thirty-eight (34.5%) patients required prone ventilation, 24 (21.8%) received non-invasive ventilation, and 42 (38.2%) patients required invasive mechanical ventilation, of whom 7 received extracorporeal membrane oxygenation as rescue therapy. Common complications among the 110 patients included secondary infection (51 [46.3%]), severe ARDS (42 [38.2%]), septic shock (18 [16.4%]), acute cardiovascular injury (13 [11.8%]), AKI (6 [5.4%]), and cardiac arrest (3 [2.7%]) (table 2). The patients who received early HFNC were less likely to have secondary infection or severe ARDS, and less likely to receive prone position ventilation and invasive mechanical ventilation than the patients who receive late HFNC.

Major laboratory markers and SOFA score were tracked from hospital admission (figure 2). Lymphocyte count was higher in patients who received early HFNC during hospitalization. Lactate dehydrogenase level did not differ between these two groups on day 3 after admission, but continued to increase in the late HFNC group and was significantly higher on day 9 after admission and onwards. Both the levels of D-dimer and C-reactive protein were significantly lower throughout the clinical course in patients who received early HFNC. Lactate concentration and SOFA score were similar between groups on day 3 after admission. Patients who received early HFNC showed lower lactate concentration on day 15 and lower SOFA score on day 9 after admission and onwards (figure 2).

**Figure 2.**
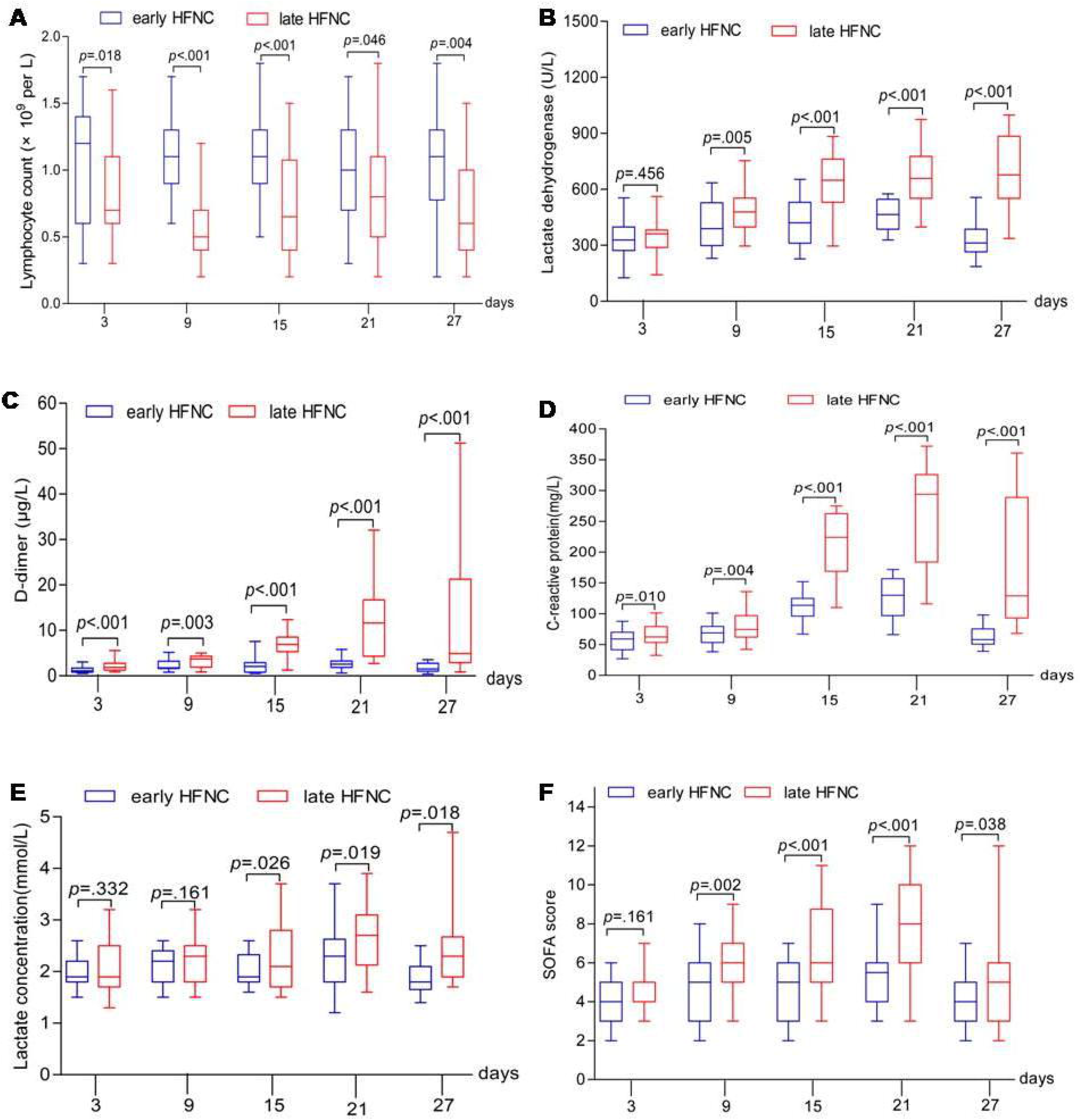
Dynamic changes in major laboratory markers and SOFA score in elderly patients with COVID-19. Figure shows temporal changes in lymphocyte (A), lactate dehydrogenase (B), D-dimer (C), C-reactive protein (D), lactate (E), and SOFA score (F) after admission. COVID-19=coronavirus disease 2019, SOFA score =Sequential Organ Failure Assessment score, HFNC=High-flow nasal cannula oxygen therapy. The horizontal lines represent the median value in each group.

Of the 110 patients who received HFNC, 40 (36.4%) patients required more than 60% FiO_2_, 23 (22.7%) patients required FiO_2_ more than 80%, and 4 (3.6%) patients required 100% FiO_2_ (table 1). All 22 patients in the late HFNC group who died during hospitalization received FiO_2_ > 60% for more than 72 hours continuously (table 3). The numbers of patients who required higher than 60% FiO_2_ and those who required more than 80% FiO_2_ were smaller in the early HFNC group. FiO_2_ and PO_2_/FiO_2_ were tracked during hospitalization. As shown in figure 3, baseline FiO_2_ and PaO_2_/FiO_2_ were similar between the two groups. The patients in the early HFNC group showed higher ratio of PaO_2_/FiO_2_ on day 3 after admission, and required lower FiO_2_ on day 6 after admission and onwards.

**Figure 3.**
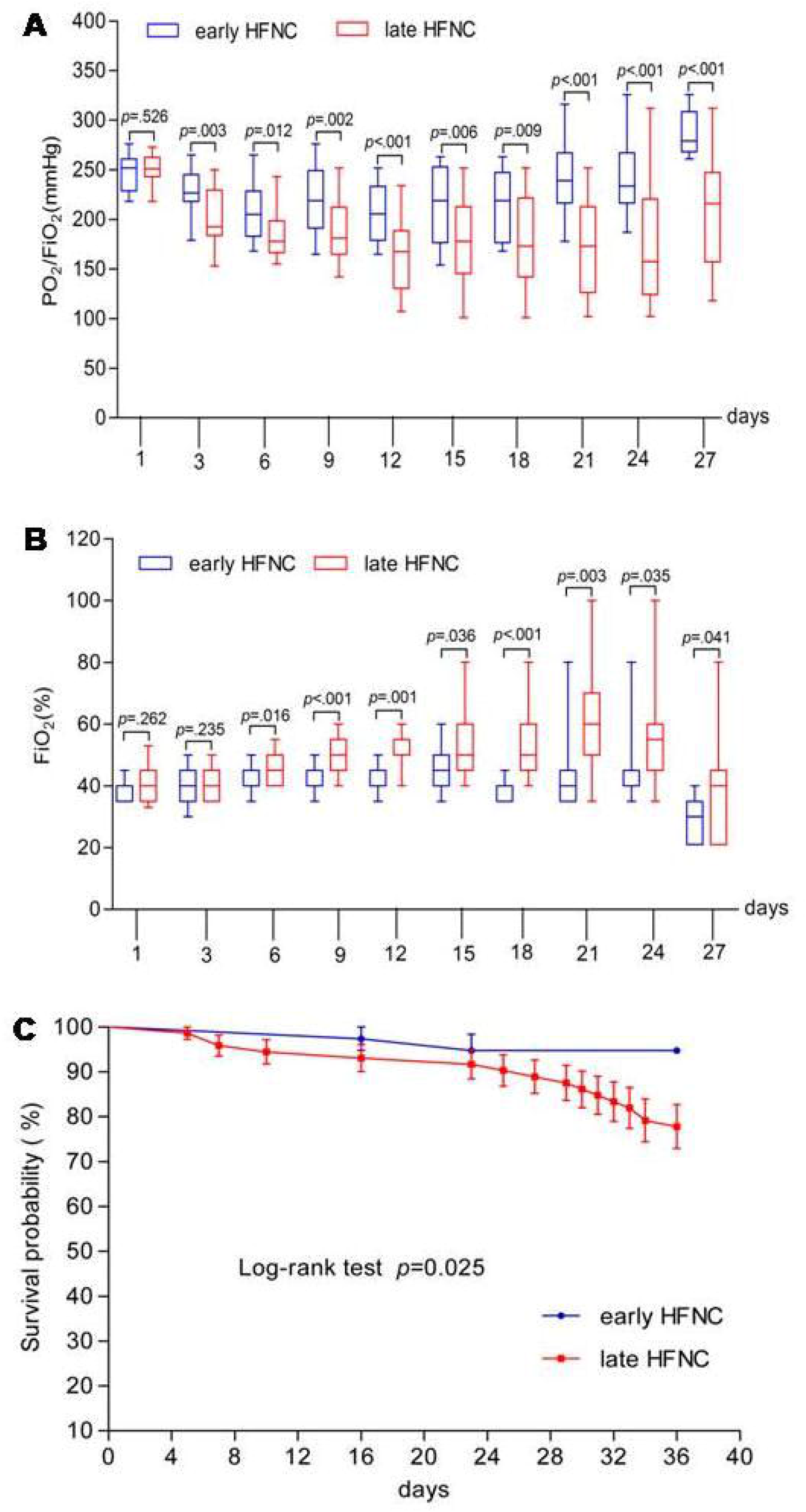
Dynamic changes of PaO2/FiO2 and FiO2 and survival probability in patients with COVID-19. Figure shows temporal changes in PaO_2_/FiO_2_ (A) and FiO_2_ (B), and survival probability (C). COVID-19=coronavirus disease 2019, PaO2/FiO2=Ratio of arterial oxygen partial pressure to fraction inspired oxygen concentration, FiO2 =fraction inspired oxygen concentration. The horizontal lines represent the median value in each group in A and B.

**Table 3.** Clinical measures of 72 elderly patients with COVID-19 who received late HFNC group

| Clinical measures | Total<br>(n=72) | Non-survivors<br>(n=22) | Survivors<br>(n=50) | *P* value<sup>a</sup> |
| --- | --- | --- | --- | --- |
| Heart rate, beats per min | 92 (68-139) | 93 (62-141) | 91 (65-132) | 0.287 |
| Systolic blood pressure on admission, mmHg | 134 (115-158) | 138 (110-163) | 130 (105-165) | 0.565 |
| Diastolic blood pressure on admission, mmHg | 75 (66-115) | 77 (63-115) | 75 (65-110) | 0.431 |
| SOFA score on admission | 4 (3-6) | 5 (3-7) | 4 (3-6) | 0.127 |
| APECHE □ score on admission | 14 (12-19) | 15 (12-19) | 14 (12-17) | 0.602 |
| Comorbidities per person | 1.4 (0.8-1.6) | 1.6 (0.9-1.7) | 1.3 (0.6-1.6) | 0.211 |
| Haemoglobin concentration on admission, g/L | 125 (97-136) | 123 (92-138) | 125 (98-143) | 0.341 |
| Lymphocyte count on admission, ×10<sup>9</sup>/L | 0.75 (0.35-1.65) | 0.73 (0.32-1.63) | 0.76 (0.39-1.71) | 0.791 |
| Platelet count on admission, ×10<sup>9</sup>/L | 165 (123-223) | 165 (121-231) | 167 (126-226) | 0.463 |
| Total bilirubin concentration on admission, μmol/L | 9.3 (6.7-15.6) | 9.6 (6.4-16.2) | 9.2 (6.5-14.3) | 0.358 |
| Serum creatinine concentration on admission, μmol/L | 78.7 (55.6-101.2) | 83.5 (70.2-102) | 71.2 (53.3-91.5) | 0.212 |
| Lactate concentration on admission, mmol/L | 1.7 (1.5-3.2) | 1.7 (1.3-3.1) | 1.6 (1.3-3.2) | 0.411 |
| Ratio of PaO<sub>2</sub> to FiO<sub>2</sub>, mmHg on admission | 173 (155-197) | 174 (132-198) | 172 (152-195) | 0.637 |
| Invasive Mechanical ventilation | 38 (52.7%) | 22 (100%) | 16 (32%) | <0.001 |
| **FiO<sub>2</sub> during hospitalization** |  |  |  |  |
| 100% more than 72 hours continuously, n | 4 (5.5%) | 4 (18.1%) | 0 | 0.011 |
| >80% more than 72 hours continuously, n | 16 (22.2%) | 12 (54.5%) | 4 (8%) | <0.001 |
| >60% more than 72 hours continuously, n | 32 (44.4%) | 22 (100%) | 12 (24%) | <0.001 |

Data are median (IQR), n (%), or n/N (%). FiO<sub>2</sub>=fraction of inspired oxygen. HFNC=high flow nasal cannula. PaO<sub>2</sub>=partial pressure of oxygen.
<sup>a</sup>*P* values indicate differences between Non-survivors and Survivors.
*P* < 0.05 was considered statistically significant.

The median time from illness onset to ICU admission was 11 days (IQR 8-14), and 12 days (IQR 11-15) to severe ARDS. The length of ICU stay was 17 days (IQR 12-22), LOS was 27 days (IQR 16-32), and duration of viral shedding after illness onset was 16 days (IQR 13-21). Patients who received early HFNC were less likely to admit to ICU, less likely to stay in ICU longer than 7 days, had less chance to develop severe ARDS and had longer time from COVID-19 onset to severe ARDS (if any). Duration of viral shedding after illness onset and length of ICU and hospital stay were shorter in the early HFNC group. A total of 86 (78.2%) patients had been discharged, and 24 (21.8%) patients had died. The mortality in late HFNC group was higher than that in early HFNC (22 [30.6%] vs. 2 [5.3%]) as shown in Table 2 and Figure 3C.

## Discussion

This report, presents the outcomes of 110 severely ill elderly COVID-19 patients who received oxygen therapy with HFNC. Mortality was lower (5.3%) in 38 patients who received HNFC treatment at the mild ARDS stage, compared to 30.6% in 72 patients in whom HNFC treatment was started at the moderate ARDS stage. The mortality rate in this study was lower than that reported from a multi-national study regarding the mortality for patients with ARDS, which was 34.9% and 40.3% respectively for those with mild or moderate ARDS.[17] An early report from China[3] and a recent report from the United States[5] reported 61.5% and 50% mortalities respectively for critically ill patients with COVID-19, while the mortality for critically ill patients aged 60 years or older was as high as 70.3%.[3]

HFNC, as an innovative and effective modality for oxygen therapy, delivers titratable oxygen up to 60 liters/minutes with heating and humidification to produce a low-level positive end-expiratory pressure and to achieve FiO_2_ as high as 95-100%.[9] HFNC has been shown to reduce the risk of requiring more advanced ventilation and relieve dyspnea better than conventional oxygen therapy and has been suggested as a first-line therapy even before making a clear diagnosis for dyspnea.[10] In our study, 10.5% patients in the early HFNC group converted to invasive mechanic ventilation, which is in contrast to the 52.7% in the late HFNC group (Table 2). These findings can be compared to other published reports. A cohort study in 17 COVID-19 patients indicated starting HFNC when PaO_2_/FiO_2_>200 reduced the need of mechanical ventilation, although the impact on mortality was not reported.[18] Starting HFNC or invasive mechanical ventilation at a relatively late stage of disease severity such as moderate to severe ARDS may prompt the physician to apply high FiO_2_. Critically ill patients with COVID-19 in the Seattle region[5] had reported 50% mortality at the time of data cut off with several patients continuing to receive mechanical ventilation in the ICU. In the study,[5] the initial median FiO_2_ on day 1 of mechanical ventilation was 90% (IQR 70-100%), and the FiO_2_ decreased to 60% (IQR 50-70%) on day 3 but no further information was provided about FiO_2_ afterwards. It is possible that the FiO_2_ had to be readjusted to higher levels due to the subsequent difficulty in reaching targeted PaO_2_ and/or PaO_2_/FiO_2_. High oxygen mediated oxidative lung damage[19] may further exacerbate oxygenation, which may paradoxically push for the need of higher FiO_2_. In addition, oxidative stress during respiratory viral infection may also exacerbate a “cytokine storm”.[20] In the late HFNC group, the required FiO_2_ increase over time we found (Figure 3) was coincident with progressive increases of D-dimer and C-reactive protein (Figure 2), indicators of inflammation that could be related to a relatively higher mortality rate in the late HFNC group.

Evidence shows that liberal oxygen therapy increases mortality without improving other outcomes and that supplemental oxygen might become unfavorable above a SpO_2_ range of 94-96%.[21] A multicenter study of critically ill patients with the Middle East Respiratory Syndrome (MERS) related to MERS-CoV infection showed that non-survivors received significantly higher FiO2 than survivors on ICU day 1.[22] Thus, despite the generally accepted normal range of PaO_2_ 80 - 100 mmHg breathing room air at sea level in healthy young adults, we took into consideration the relatively lower reference values for PaO_2_ in the elderly compared to young adults as well as gender differences.[23’ 24] Previous studies showed that in elders over 70 years old the normal PaO_2_ for men was 77 mmHg (SD. 9.1; and lower limit of normal at 62mmHg), while PaO_2_ for women was 73.5 mmHg (SD. 8.4; lower limit of normal at 59.6mmHg), [23]and normal reference values reduce with age.[24] In practice, we estimated the acceptable normal values of PaO_2_ using the formula: normal PaO_2_ at sea level (in mmHg) = 100 minus the number of years over ago 40, as proposed.[25] For SpO_2_, we recommended 93% for men and 92% for women as the lower limit of normal. We also recommended SpO_2_ 95% or 96% as the highest target value, which is generally in keeping with the recommendation by Chinese CDC and the recently published guideline recommending of no higher than 96%.[26] However, accuracy of SpO_2_ readings may be affected by factors such as low perfusion and the use of vasodilator,[27] so target values of SpO_2_ were set at the discretion of treating physician, and arterial blood gas analysis was used to adjust treatments (e.g., FiO_2_ and/or flow rate).

In our study, FiO_2_ values were maintained significantly lower in the early HFNC group (Figure 2). Post-hoc subgroup analysis in the late HFNC group revealed that FiO_2_ of survivors was significantly lower than that of the non-survivors (table 3), and initial targeted SpO_2_ was also relatively higher in the non-survivor subgroup (data not shown). In the current study, all the baseline characteristics and laboratory values were comparable between early and late HFNC groups.

There is evidence to show that airborne transmission with HFNC is minimal[11’ 28] and that risk of hospital-acquired infection did not increase with the use of HFNC provided there is good mask fitting.[29] However, the safety of HFNV in these patients is controversial given SARS-CoV-2 virus is highly contagious.[30] Because of risks, all staff in ward or ICU care settings are strongly recommended/required to wear a disposable surgical cap, medical protective mask (N95), disposable medical protective uniform and disposable gloves with full-face respiratory protective devices when performing procedures like tracheal intubation.[31]

In conclusion, the application of HFNC in elderly patients (≥ 65 years) with COVID-19, especially when used with conservative oxygen delivery, may prove to be a promising treatment modality for critically ill patients with acute ARDS in general, and of critically ill elderly COVID-19 patients in particular, although larger scale prospective studies are needed to confirm its effectiveness. Our current study provides evidence that application of HFNC earlier during the mild stage of ARDS may be associated with reduced need for mechanic ventilation and mortality in critically ill elderly patients with COVID-19 pneumonia. The fact that early application of HFNC was associated with shorter time duration of SARS-CoV-2 viral shedding may be of significance in reducing transmission.

### Clinical Perspectives

- High-flow nasal cannula therapy (HFNC) is a major oxygen supporting therapy for severely ill patients, and is recommended for use in COVID-19 patients. However, study is lacking regarding the optimal timing of high-flow nasal cannula (HFNC) application among critically ill elderly COVID-19 patients. We hypothesized that early application of HFNC for oxygen delivery in severely ill COVID-19 patients may facilitate patient recovery and reduce mortality.
- In this retrospective, multicenter cohort study involving 110 elderly patients with laboratory-confirmed COVID-19, prognosis was much better in 38 patients who received HFNC when 200 mmHg < PaO2/FiO2 ≤ 300 mmHg, compared to 72 patients who received HFNC treatment when their 100 mmHg< PaO2/FiO2 ≤ 200 mmHg. Early application of HFNC was associated shorter lengths of ICU and hospital stay and reduced mortality.
- HFNC should be considered early in treating elderly patients with COVID-19.

### Contributors

LD and ZX had the idea for the study. ZX, LD and JT designed the study and have full access to all data in the study and take responsibility for the integrity of the data and the accuracy of the data analysis. SL, LZ, DL, CH, DZ, ZW, XS, and YZ collected the data. FJ, SL and DL performed data analysis. DAL, Z-YX, SW, CWC, HL and RLA participated in discussion/data interpretation. SL, FJ, DL and ZX drafted the manuscript. RLA, HL and ZX revised the final manuscript.

### Data sharing

With the permission of the corresponding authors, the participant data without names and identifiers can be provided, but not the study protocol and statistical analysis plan. The data will be available for others to request after publication of study findings. The research team will provide an email address for communication once the data can be made public. The corresponding authors have the right to decide whether or not to share the data based on research objectives and plan provided.

## Acknowledgements

The authors’ work was supported by the grants from National Natural Science Foundation of China (NSFC 813000674, 81670770, 81970247). We acknowledge all health-care workers involved in the diagnosis and treatment of patients in Wuhan. We thank the patients and their families for providing requested data and information.

